# Effects of Prenatal Substance Exposure on Longitudinal Tri-Ponderal Mass Index Trajectories from Pre- to Early Adolescence in the ABCD study

**DOI:** 10.1101/2024.11.23.24317835

**Authors:** Ru Li, Isabella Mariani Wigley, Ilkka Suuronen, Ashmeet Jolly, Jetro J. Tuulari

## Abstract

**Objectives:** The long-term relationship between prenatal substance exposure (PSE) and obesity development remains inconclusive and poorly understood. This study aimed to explore the heterogeneity in adiposity developmental trajectories from pre- to early adolescence and investigate the influence of PSE on these patterns.

**Methods:** Five waves of data from 7 881 children enrolled in the Adolescent Brain Cognitive Development (ABCD) Study (Release 5.1) were analyzed. Tri-Ponderal Mass Index (TMI) was used to assess adiposity levels. PSE (e.g., tobacco, alcohol, caffeine, and marijuana) was collected via maternal self-report. Latent growth mixture modeling was conducted to identify TMI trajectories, followed by multinomial logistic regression to examine the role of PSE in TMI profiles, controlling for various factors.

**Results:** Three trajectories were identified: *Stable TMI* (86.6%), *Increasing TMI* (12.5%), and *Decreasing TMI* (0.9%). The risk of exhibiting an *Increasing TMI* was associated with prenatal exposure to tobacco (β = 1.53, 95% CI = 1.26–1.86, p < .001) and caffeine (daily use: β = 1.39, 95% CI = 1.16–1.68, p < .001; weekly use: β = 1.38, 95% CI = 1.13–1.69, p < .05), with dose-dependent effects. Notably, tobacco exposure both before (β = 1.55, 95% CI = 1.27–1.89, p < .001) and after awareness of pregnancy (β = 1.51, 95% CI = 1.10–2.08, p < .05) contributed to this risk, with no significant benefit from maternal cessation after pregnancy awareness. Multiple PSE substantially elevated the risk of increasing adiposity (β = 1.70, 95% CI = 1.27–2.27, p < .001).

**Conclusions:** Obesity risk can emerge long before adolescence, even during prenatal development. The findings regarding the long-term influence of prenatal substance exposure on adiposity development during adolescence highlight the importance of preconception and prenatal health interventions to mitigate the risk of obesity in offspring.

## Introduction

Over the past four decades, obesity rates doubled in more than 70 countries and consistently increased in most others [1]. The prevalence of obesity among children and adolescents has been growing rapidly [2]. The rising rates of obesity in children not only pose immediate health concerns but also increase the risk of obesity persisting into adulthood, where the long-term effects can be even more detrimental [3–5]. Therefore, understanding the factors that influence the developmental trajectory of adiposity during adolescence is essential for effective prevention and intervention strategies.

The etiology of obesity is complex, involving interactions between genetic, environmental, and behavioral factors. Maternal substance use during pregnancy, including tobacco, alcohol, caffeine, and marijuana, has been associated with various adverse offspring outcomes [6,7]. The Developmental Origins of Health and Disease hypothesis posits that environmental exposures during critical periods of development can have long-lasting effects on health and disease risk [8]. This notion underscores the importance of considering prenatal substance exposure (PSE) when exploring the roots of obesity. For example, maternal smoking was suggested to potentially affect fetal growth and development due to altered placental function, oxidative stress, and epigenetic modifications [9].

Despite the accumulating evidence on the role of prenatal substance exposure in influencing offspring health outcomes, the long-term effects of these exposures on adiposity development during adolescence remain poorly understood. Most studies to date have focused on single-time-point assessments of obesity, which may not adequately capture the dynamic changes in body composition over time, resulting in mixed findings. For instance, the effects of prenatal alcohol exposure on adiposity have shown differential results across studies [10–12]. Additionally, research on prenatal caffeine and marijuana exposure in relation to offspring adiposity is relatively scarce and inconclusive [13–15].

The inconsistency may also be attributed to the amount of use, changes in exposure status (e.g., from exposed to unexposed), and postnatal substance exposures, which can have varying effects on fetal development and subsequent health outcomes [16,17]. Furthermore, most previous research has focused on the effect of single substance, neglecting the potential cumulative effects of multiple exposures, which is especially relevant since many pregnant women who use substances often use more than one type [18].

Conventionally, body mass index (BMI) has been widely used to assess obesity. However, during adolescence, hormonal shifts and growth spurts can affect body mass in ways BMI may not fully capture, particularly in distinguishing between lean and fat mass. Consequently, the tri-ponderal mass index (TMI), calculated as weight divided by height cubed, has been proposed to be a better metric for evaluating adiposity in adolescents [19,20]. TMI accounts for the changing body proportions during growth and has shown superior performance in estimating body fat percentage compared to BMI, especially during puberty. It is considered an accurate and stable indicator for children and, unlike BMI, is not dependent on national weight distributions, making it ideal for international comparisons. Additionally, previous research using models that assume no within-group variances typically revealed similar growth patterns across three or four BMI trajectory groups [21,22]. Nevertheless, given the complex dynamic changes during early adolescence, we expected heterogeneity within groups.

The primary objective of this study was to explore how prenatal substance exposure influences adiposity trajectories over time, with three specific aims: 1) to identify distinct TMI trajectories over four years in children who were predominantly between the ages of 9 and 14 years (from pre-adolescence to early adolescence); 2) to investigate relationships between these trajectories and prenatal exposure to tobacco, alcohol, caffeine, and marijuana, expecting varying effects based on type and amount of exposure; and 3) to assess the influence of exposure timing (before and after awareness of pregnancy), exposure status change, and the number of substances used, hypothesizing that multiple exposures would have a stronger impact than single-substance exposure.

## Methods

### Study design and participants

The ABCD Study (Release 5.1) included data from 11 868 children aged 9 to 10 years old enrolled between 2016 and 2018 (https://abcdstudy.org). Detailed information about the study design, recruitment strategy, sample selection procedure, and assessments is available in the original source [23]. This release contains data from a subset of participants due to varying numbers of missed visits for each event and the ongoing 4-year follow-up at the time the data were frozen. For the current study, individuals with complete anthropometric measurement data were included, which is available for 11 856 participants at baseline (Y0), 11 137 at the 1-year follow-up (Y1), 9 151 at the 2-year follow-up (Y2), 3 777 at the 3-year follow-up (Y3), and 4 178 at the 4-year follow-up (Y4). The study adhered to the guidelines proposed by the Strengthening the Reporting of Observational Studies in Epidemiology (STROBE) [24].

The sample for this study was obtained based on multiple exclusion criteria, as outlined in the flowchart (**Figure 1**) and detailed in the Supplementary Material Section 1. The final sample consisted of 7 881 children. Of these, 7 877 children were available at Y0, 7 690 at Y1, 6 318 at Y2, 2 596 at Y3, and 2 860 at Y4.

**Figure 1.**
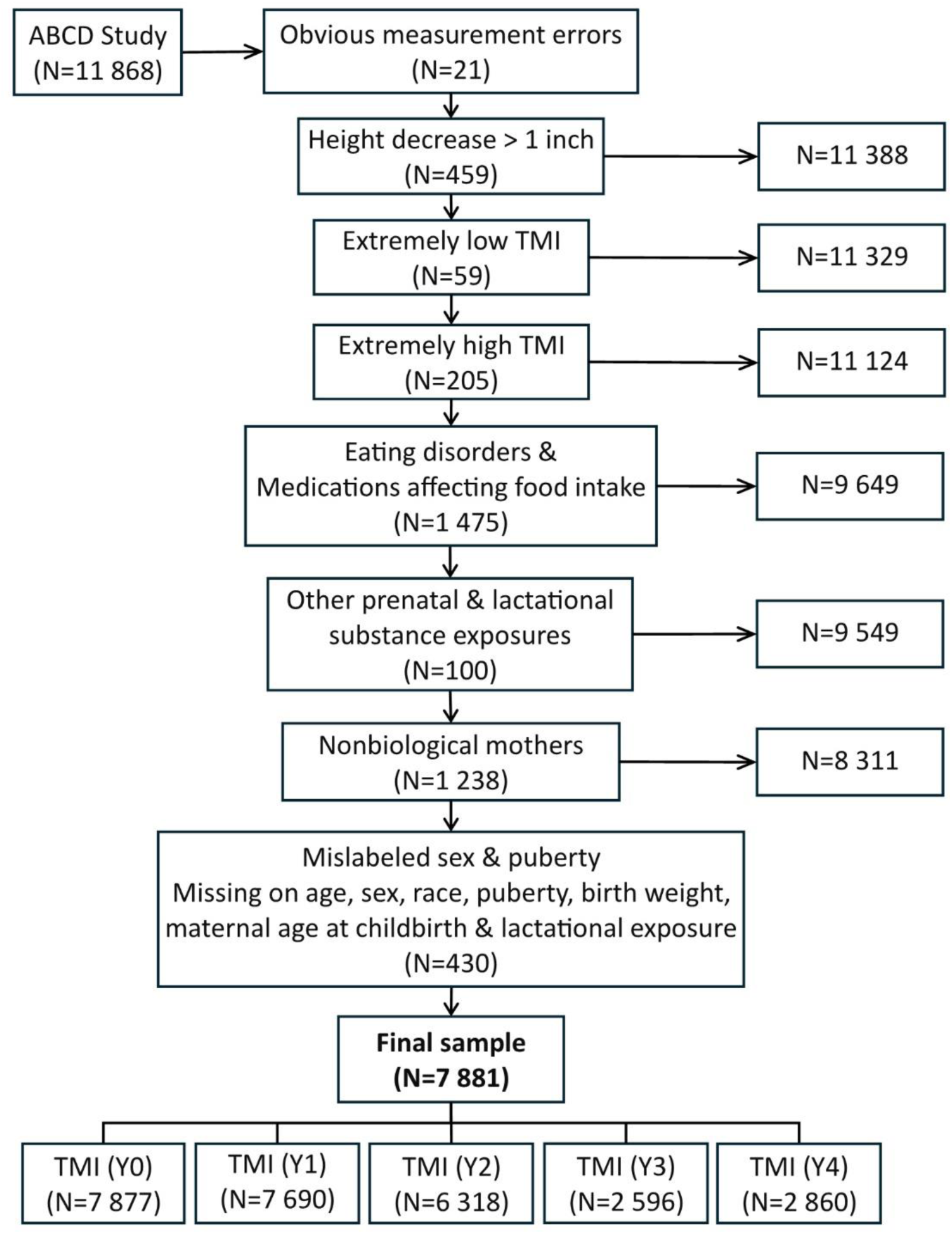
Flowchart of sample inclusion and exclusion criteria

The data from the ABCD Study are held in the NIMH Data Archive. The ABCD Study is responsible for obtaining participant consent and assent, with protocols approved by a centralized Institutional Review Board at the University of California, San Diego [25].

## Measures

### Tri-ponderal mass index (TMI)

The average height (in.) and weight (lbs.), measured in light clothing and stocking feet at each wave, were converted to TMI using the formula weight (kg) / height (m)^3^. In this study, the conversion was (weight (lbs.) / 2.205) / (height (in.) / 39.370)^3^.

### Prenatal exposures to substances

At the baseline of the ABCD study, the Developmental History Questionnaire (DHQ) was used to inquire about the use of specific substances during pregnancy, including tobacco, alcohol, caffeine, marijuana, and other substances such as cocaine, heroin, and oxycodone [26]. Maternal reports indicated prenatal exposure to tobacco, alcohol, and marijuana, recorded as any use before and after pregnancy awareness (yes or no). Prenatal caffeine exposure was reported without specific timing. Response options for caffeine included: nonuse, daily use (at least once a day), weekly use (less than once a day but more than once a week), and monthly use (less than once a week).

The amount of exposure was measured by asking about the daily frequency of smoking and marijuana use, the maximum number of alcoholic drinks consumed in one sitting, average weekly alcohol intake, and weekly caffeine consumption (calculated from daily intake × 7 and monthly intake / 4.33).

Moreover, if any prenatal use of tobacco, alcohol, or marijuana played a role, the exposures before and after awareness of pregnancy were compared. These were further divided into non-use (no consumption before or after knowing about the pregnancy), abstain (consumption before but not after knowing about the pregnancy), and consistent use (consumption both before and after knowing about the pregnancy). In addition, exposure levels were classified into 4 categories: non-exposure, mono-exposure (using one substance), dual-exposure (using two substances), and poly-exposure (using more than two substances).

### Background information

Data on past or current eating disorders (Y2 and Y4) and medications affecting food intake (across all waves) were collected. Lactational substance exposure and childhood substance use were treated as confounders. Demographic variables—including age, sex, race, pubertal stage, birth weight, maternal age at childbirth, and maternal education (as a socioeconomic indicator)—were controlled for in the analysis (see Supplementary Material Section 2 for details).

### Analytic strategy

To better capture TMI trajectories, background variables were first adjusted using multiple regression to obtain predicted TMI values. Adjusted TMI Z scores, calculated as Z = (V - V0) / RMSE (where V is the observed TMI, V0 the predicted value, and RMSE the root mean square error), were then used for modeling.

We implemented the modeling primarily based on established guidelines for longitudinal latent variable mixture modeling [27,28]. In brief, a single-class growth curve model was first performed to assess overall TMI development. Latent growth mixture modeling (LGMM) was then conducted to identify subpopulations with different TMI growth patterns, testing 2-to 5-class solutions. As a potential alternative model, latent class growth models (LCGM) assuming no within-class variances were also performed. The modeling employed robust maximum likelihood estimation for mixture model analysis, using 500 random starts and 20 initial iterations to optimize global solutions and stabilize parameter estimation. Model selection was based on model fit indices and interpretability. Details on the missing data mechanism, modeling procedures, and model fit indices for model selection are provided in the Supplementary Material.

Subsequently, zero-order relationships between all variables and the identified TMI trajectory groups were examined. Chi-square tests were used for categorical variables. Either one-way ANOVA (S-N-K post hoc test) or Kruskal-Wallis test was used for group comparisons on continuous variables, depending on the homogeneity or heterogeneity of variance. Multinomial logistic regression analysis was conducted to evaluate the influence of prenatal substance exposure on TMI trajectories, controlling for demographics and relevant confounders.

Descriptive statistics and attrition analysis were performed in IBM SPSS 29. All modeling was conducted using Mplus 8.3 software [29]. Additional statistical analyses and figure generation were completed in R (version 4.4.1).

## Results

### Descriptive statistics and attrition analysis

**Table 1** presents the descriptive statistics of sample characteristics and prenatal substance exposures across five waves, including information on missing data, as we did not exclude cases with missing values on the focal predictive variables. The final sample of this study consists of 3 978 boys and 3 903 girls, with 99.5% being 9 or 10 years old at baseline.

**Table 1.** Characteristics and prenatal substance exposures of sample available at each wave.

|  | Y0<br>(N = 7877) | Y1<br>(N = 7690) | Y2<br>(N = 6318) | Y3<br>(N = 2596) | Y4<br>(N = 2860) | *P* |
| --- | --- | --- | --- | --- | --- | --- |
| *N (%) or M (SD)* |  |  |  |  |  |  |
| **Sex** |  |  |  |  |  | .820 |
| *Males* | 3976 (50.5%) | 3874 (50.4%) | 3227 (51.1%) | 1328 (51.2%) | 1469 (51.4%) |  |
| *Females* | 3901 (49.5%) | 3816 (49.6%) | 3091 (48.9%) | 1268 (48.8%) | 1391 (48.6%) |  |
| **Race** |  |  |  |  |  | < .001 |
| *White* | 4182 (53.1%)<sup>a</sup> | 4119 (53.6%)<sup>ab</sup> | 3521 (55.7%)<sup>bc</sup> | 1421 (54.7%)<sup>abc</sup> | 1641 (57.4%)<sup>c</sup> |  |
| *Black* | 1116 (14.2%)<sup>a</sup> | 1064 (13.8%)<sup>a</sup> | 829 (13.1%)<sup>a</sup> | 365 (14.1%)<sup>a</sup> | 308 (10.8%)<sup>b</sup> |  |
| *Hispanic* | 1647 (20.9%) | 1592 (20.7%) | 1239 (19.6%) | 493 (19.0%) | 584 (20.4%) |  |
| *Asian* | 151 (1.9%) | 149 (1.9%) | 113 (1.8%) | 40 (1.5%) | 52 (1.8%) |  |
| *Other* | 781 (9.9%) | 766 (10.0%) | 616 (9.7%) | 277 (10.7%) | 275 (9.6%) |  |
| Age (baseline) | 9.48 (0.51) | 9.48 (0.51) | 9.48 (0.51) | 9.46 (0.50) | 9.51 (0.50) | .002<sup>1</sup> |
| Puberty | 2.56 (0.74) | 2.55 (0.74) | 2.57 (0.73) | 2.57 (0.74) | 2.68 (0.70) | < .001<sup>2</sup> |
| BW (lbs.) | 6.58 (1.46) | 6.58 (1.46) | 6.55 (1.47) | 6.53 (1.48) | 6.53 (1.50) | .279<sup>2</sup> |
| ME | 16.67 (2.73) | 16.70 (2.72) | 16.76 (2.64) | 16.73 (2.64) | 16.85 (2.62) | .069<sup>1</sup> |
| MAC | 29.52 (6.11) | 29.57 (6.07) | 29.58 (6.07) | 29.49 (5.94) | 29.77 (5.99) | .393<sup>2</sup> |
| **PSE** |  |  |  |  |  |  |
| **Tobacco** | 935 (12.0%) | 906 (11.9%) | 745 (11.9%) | 330 (12.8%) | 306 (10.8%) | .252 |
| Missingness | 58 (0.7%) | 58 (0.8%) | 49 (0.8%) | 15 (0.6%) | 21 (0.7%) |  |
| **Alcohol** | 1906 (25.2%) | 1872 (25.3%) | 1537 (25.3%) | 623 (25.0%) | 703 (25.5%) | .991 |
| Missingness | 304 (3.9%) | 301 (3.9%) | 251 (4.0%) | 101 (3.9%) | 108 (3.8%) |  |
| **Caffeine** |  |  |  |  |  | .857 |
| *Daily* | 1824 (24.4%) | 1779 (24.4%) | 1503 (25.0%) | 583 (23.5%) | 689 (25.2%) |  |
| *Weekly* | 1480 (19.8%) | 1449 (19.9%) | 1198 (19.9%) | 507 (20.5%) | 559 (20.5%) |  |
| *Monthly* | 1192 (16.0%) | 1167 (16.0%) | 974 (16.2%) | 426 (17.2%) | 424 (15.5%) |  |
| Missingness | 406 (5.2%) | 394 (5.1%) | 308 (4.9%) | 117 (4.5%) | 127 (4.4%) |  |
| **Marijuana** | 394 (5.0%)<sup>a</sup> | 382 (5.0%)<sup>a</sup> | 301 (4.8%)<sup>ab</sup> | 125 (4.9%)<sup>ab</sup> | 103 (3.6%)<sup>b</sup> | .037 |
| Missingness | 72 (0.9%) | 71 (0.9%) | 55 (0.9%) | 23 (0.9%) | 27 (0.9%) |  |

BW = Birth weight. ME = Maternal education levels. MAC = Maternal age at childbirth. PSE = Prenatal substance exposure. Focal variables are bolded.
Groups sharing the same superscript = no significant differences.
<sup>1</sup>Kruskal-Wallis test
<sup>2</sup>One-way ANOVA (S-N-K post hoc test)

At Y4, the proportion of Black children was lower, and participants appeared to have higher puberty levels compared to earlier waves. No significant differences in sex, birth weight, maternal education, or maternal age at childbirth were observed across the five waves, indicating consistent sample characteristics. Regarding prenatal substance exposure, marijuana exposure was lower at Y4 than at Y0 and Y1.

### Growth curve modeling

Both the linear and quadratic models demonstrated good fit to the data, with the quadratic model showing a better fit (see Section 5.1 in Supplementary Material for details). The overall TMI level remained stable over time, but there were significant variances in baseline levels and slopes. Further, a good fit of the model with equal error variances confirmed measurement reliability, supporting the robustness of subsequent modeling to identify distinct growth patterns among subgroups. The single-class model is illustrated in Supplementary **Figure S1**.

### Latent growth mixture modeling (LGMM)

Detailed descriptions and model fit indices are provided in Section 5.2 of the Supplementary Material. In brief, the quadratic model showed a better model fit than the linear model. Model fit indices did not indicate a significantly superior fit beyond the 3-class model. Moreover, compared to the 3-class model, the 4-class and 5-class models exhibited a more skewed class distribution. Finally, the 3-class quadratic model with freely estimated intercepts across classes was preferred for its better fit and interpretability.

Compared to the model assuming within-class invariances (i.e., LCGM), the LGMM provided a better fit to the data, suggesting it to be the optimal approach for modeling the TMI trajectories in this study. **Figure 2** depicts the LGMM estimated trajectories of TMI by most likely latent classes and the observed individual trajectories within each latent class (or group). The groups were named based on their development patterns. For example, the first group demonstrated consistently stable development of TMI over time and was thus named “*Stable TMI*” (N = 6 827). Likewise, the second and third groups were named “*Increasing TMI*” (N = 987) and “*Decreasing TMI*” (N = 67), respectively. Random effect parameters of the LGMM are shown in **Table 2**.

**Figure 2.**
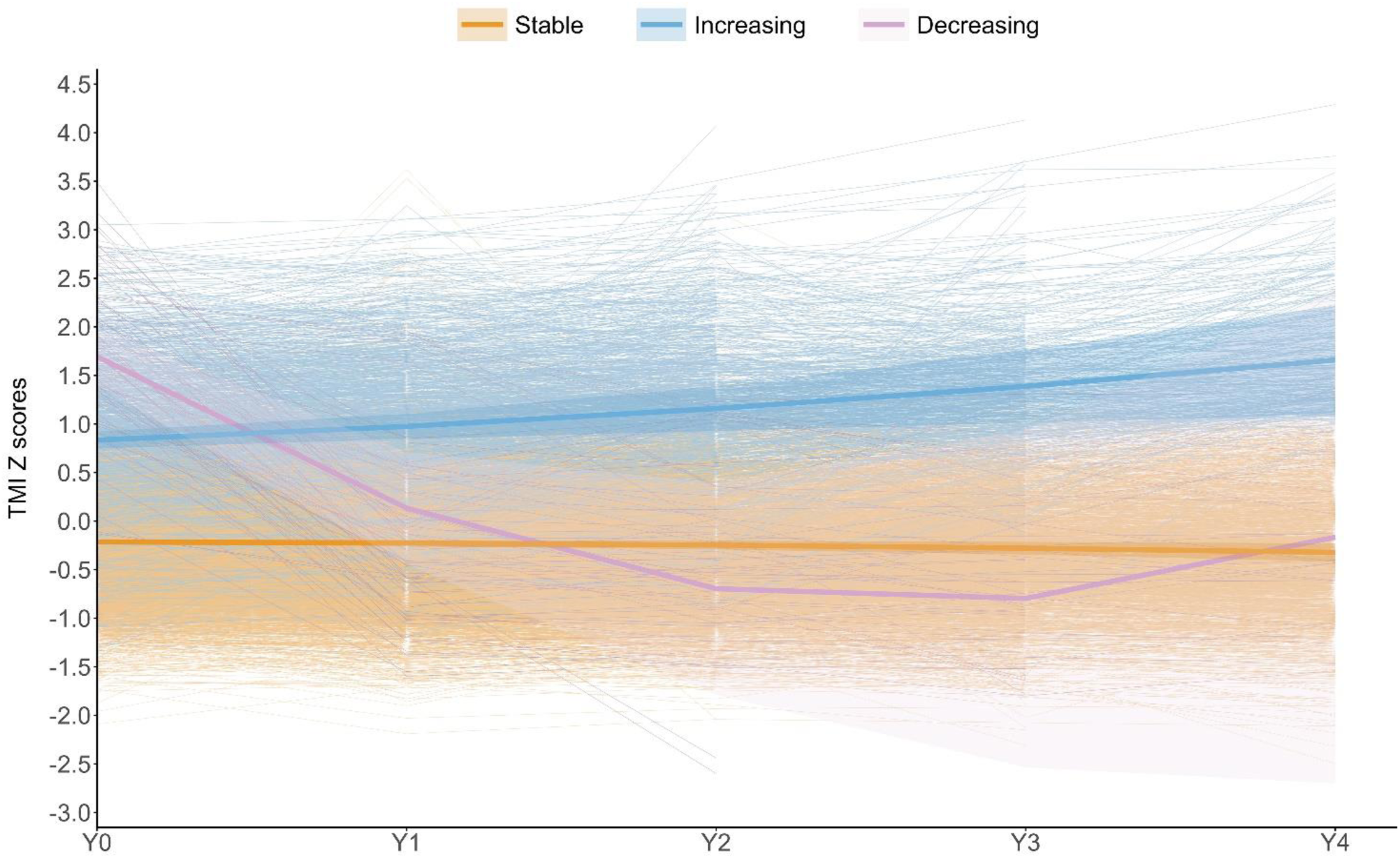
TMI trajectory patterns based on latent growth mixture modeling (shaded bands: 95% confidence intervals), with observed individual trajectories within each group

**Table 2.** Estimate and standard error (SE) of mean levels (L), slopes (S: linear; Q: quadratic), variances, and residual variances.

|  | Estimate | SE | P-value |
| --- | --- | --- | --- |
| Latent Classes |  |  |  |
| **Stable TMI** |  |  |  |
| *L* | -0.214 | 0.014 | < .001 |
| *S* | -0.005 | 0.008 | .470 |
| *Q* | -0.005 | 0.002 | .004 |
| **Increasing TMI** |  |  |  |
| *L* | 0.835 | 0.044 | < .001 |
| *S* | 0.119 | 0.048 | .013 |
| *Q* | 0.022 | 0.013 | .098 |
| **Decreasing TMI** |  |  |  |
| *L* | 1.693 | 0.151 | < .001 |
| *S* | -1.925 | 0.248 | < .001 |
| *Q* | 0.365 | 0.051 | < .001 |
| Class-Specific Variances |  |  |  |
| L (**Stable TMI**) | 0.269 | 0.016 | < .001 |
| L (**Increasing TMI**) | 0.538 | 0.038 | < .001 |
| L (**Decreasing TMI**) | 0.427 | 0.074 | < .001 |
| Fixed Variances |  |  |  |
| S | 0.018 | 0.003 | < .001 |
| Q | 0.042 | 0.009 | < .001 |
| Residual Variances |  |  |  |
| *Y0* | 0.025 | 0.006 | 0.026 |
| *Y1* | 0.100 | 0.006 | < .001 |
| *Y2* | 0.068 | 0.009 | < .001 |
| *Y3* | 0.151 | 0.017 | < .001 |
| *Y4* | 0.066 | 0.031 | 0.019 |

### Sensitivity analysis

To assess modeling robustness, three models with sample sizes ranging from 2 596 to 11 124 subjects were compared. The trajectory patterns were largely consistent, supporting FIML reliability and the model’s robustness against biases from missing data and sample exclusions (see Supplementary Material Section 6).

### Group comparisons for background variables between TMI trajectory groups

The *Stable TMI* group had more White children and fewer Black and Hispanic children compared to other groups. Children with *Decreasing TMI* had higher birth weights, while those in the *Stable TMI* group had higher maternal education levels. There were more children with tobacco exposure during breastfeeding in the *Increasing TMI* group, while lactational caffeine exposure was more prevalent in the *Stable TMI* group. Additionally, prenatal marijuana exposure was more prevalent in the *Increasing TMI* group (**Table 3**).

**Table 3.** Group comparisons for variables between TMI trajectory groups.

|  | Stable TMI | Increasing TMI | Decreasing TMI | *P* |
| --- | --- | --- | --- | --- |
|  | *N (%) or M (SD)* |  |  |  |
| Sex |  |  |  | .274 |
| Males | 3427 (50.2%) | 512 (51.9%) | 39 (58.2%) |  |
| Females | 3400 (49.8%) | 475 (48.1%) | 28 (41.8%) |  |
| Race |  |  |  | <.001 |
| White | 3720 (54.5%)<sup>a</sup> | 440 (44.6%) | 24 (35.8%) |  |
| Black | 910 (13.3%)<sup>a</sup> | 193 (19.6%) | 14 (20.9%) |  |
| Hispanic | 1392 (20.4%)<sup>a</sup> | 240 (24.3%) | 16 (23.9%) |  |
| Asian | 133 (1.9%) | 15 (1.5%) | 3 (4.5%) |  |
| Other | 672 (9.8%) | 99 (10.0%) | 10 (14.9%) |  |
| Age (baseline) | 9.47 (0.51) | 9.49 (0.50) | 9.54 (0.50) | .467<sup>1</sup> |
| Puberty | 2.55 (0.73) | 2.61 (0.79) | 2.69 (0.73) | <.001<sup>2</sup> |
| Birth Weight (lbs.) | 6.56 (1.46) | 6.65 (1.48) | 7.06 (1.22)<sup>a</sup> | <.001<sup>2</sup> |
| Maternal Education | 16.77 (2.71)<sup>a</sup> | 16.02 (2.76) | 15.76 (3.05) | <.001<sup>2</sup> |
| Maternal Age at Childbirth | 29.61 (6.11) | 28.93 (6.02) | 29.22 (6.68) | <.001<sup>2</sup> |
| **Childhood Substance Use** |  |  |  |  |
| Tobacco | 196 (2.9%) | 31 (3.1%) | 4 (6.0%) | .299 |
| Alcohol | 2280 (33.4%)<sup>a</sup> | 293 (29.7%)<sup>b</sup> | 17 (25.4%) | .029 |
| Caffeine | 5056 (74.1%)<sup>a</sup> | 780 (79.0%)<sup>b</sup> | 57 (85.1%)<sup>b</sup> | <.001 |
| **Early Substance Exposure** |  |  |  |  |
| Tobacco |  |  |  |  |
| Lactational | 93 (1.4%) | 27 (2.7%)<sup>a</sup> | 1 (1.5%) | .008 |
| Prenatal (N = 7823) | 747 (11.0%) | 178 (18.1%)<sup>a</sup> | 10 (14.9%) | <.001 |
| Alcohol |  |  |  |  |
| Lactational | 591 (8.7%) | 51 (5.2%)<sup>a</sup> | 5 (7.5%) | <.001 |
| Prenatal (N = 7577) | 1670 (25.5%) | 215 (22.5%) | 21 (31.3%) | .069 |
| Caffeine |  |  |  |  |
| Lactational | 2146 (31.4%)<sup>a</sup> | 248 (25.1%) | 15 (22.4%) | <.001 |
| Prenatal (N = 7475) |  |  |  | .034 |
| *Daily* | 1559 (24.1%) | 256 (27.5%)<sup>a</sup> | 10 (15.9%) |  |
| *Weekly* | 1271 (19.6%) | 198 (21.3%) | 11 (17.5%) |  |
| *Monthly* | 1053 (16.2%) | 129 (13.9%) | 10 (15.9%) |  |
| Marijuana |  |  |  |  |
| Lactational | 46 (0.7%) | 16 (1.6%)<sup>a</sup> | 0 (0.0%) | .014 |
| Prenatal (N = 7809) | 325 (4.8%)<sup>a</sup> | 63 (6.5%) | 6 (9.0%) | .030 |
| **Exposure Level** |  |  |  | <.001 |
| Mono-exposure | 2702 (43.4%) | 386 (42.7%) | 20 (31.7%) |  |
| Dual-exposure | 1211 (19.4%) | 168 (18.6%) | 18 (28.6%) |  |
| Poly-exposure | 354 (5.7%) | 84 (9.3%)<sup>a</sup> | 3 (4.8%) |  |

Values sharing the same superscript indicate no significant differences.
<sup>1</sup>Kruskal-Wallis test
<sup>2</sup>One-way ANOVA (S-N-K post hoc test)

### Association of PSE with single-time-point TMI and TMI trajectories

In single-time-point analyses, TMI was positively correlated with prenatal tobacco and marijuana exposure, negatively correlated with alcohol exposure, and uncorrelated with caffeine exposure at any time point (**Figure S3**). Prenatal tobacco exposure had a lower prevalence in the *Stable TMI* group. In contrast to lactational caffeine exposure, daily prenatal caffeine use was more common in the *Increasing TMI* group (**Table 3**). Additionally, poly-exposure was also more prevalent in the *Increasing TMI* group.

**Figure 3** illustrates the forest plot of odds ratios (ORs) and 95% confidence intervals (CIs), reflecting the effects of prenatal substance exposures on TMI trajectories, controlling for demographic and confounding factors. Children with prenatal tobacco exposure and those with daily or weekly caffeine consumption were more likely to have *Increasing TMI*, with ORs of 1.53, 1.39, and 1.38, respectively, compared to the *Stable TMI* group (upper part of **Figure 3**). There was a trend towards a link between prenatal alcohol exposure and *Decreasing TMI* (p = .060). Higher daily tobacco use (OR = 1.06) and weekly caffeine use (OR = 1.01) by mothers were associated with a greater likelihood of *Increasing TMI* (middle part of **Figure 3**). Both tobacco use before (OR = 1.55) and after learning of pregnancy (OR = 1.51) were significant predictors of *Increasing TMI*. Notably, the significant risk remained for mothers who quit smoking after becoming aware of their pregnancy (OR = 1.47). Multiple PSE appeared to elevate the risk (OR = 1.70) of experiencing *Increasing TMI* (lower part of **Figure 3**). Additionally, compared to the *Decreasing TMI* group, only daily prenatal exposure to caffeine significantly increased the likelihood of children developing *Increasing TMI* (OR = 2.62, 95% CI: 1.21–5.65, p = .014).

**Figure 3.**
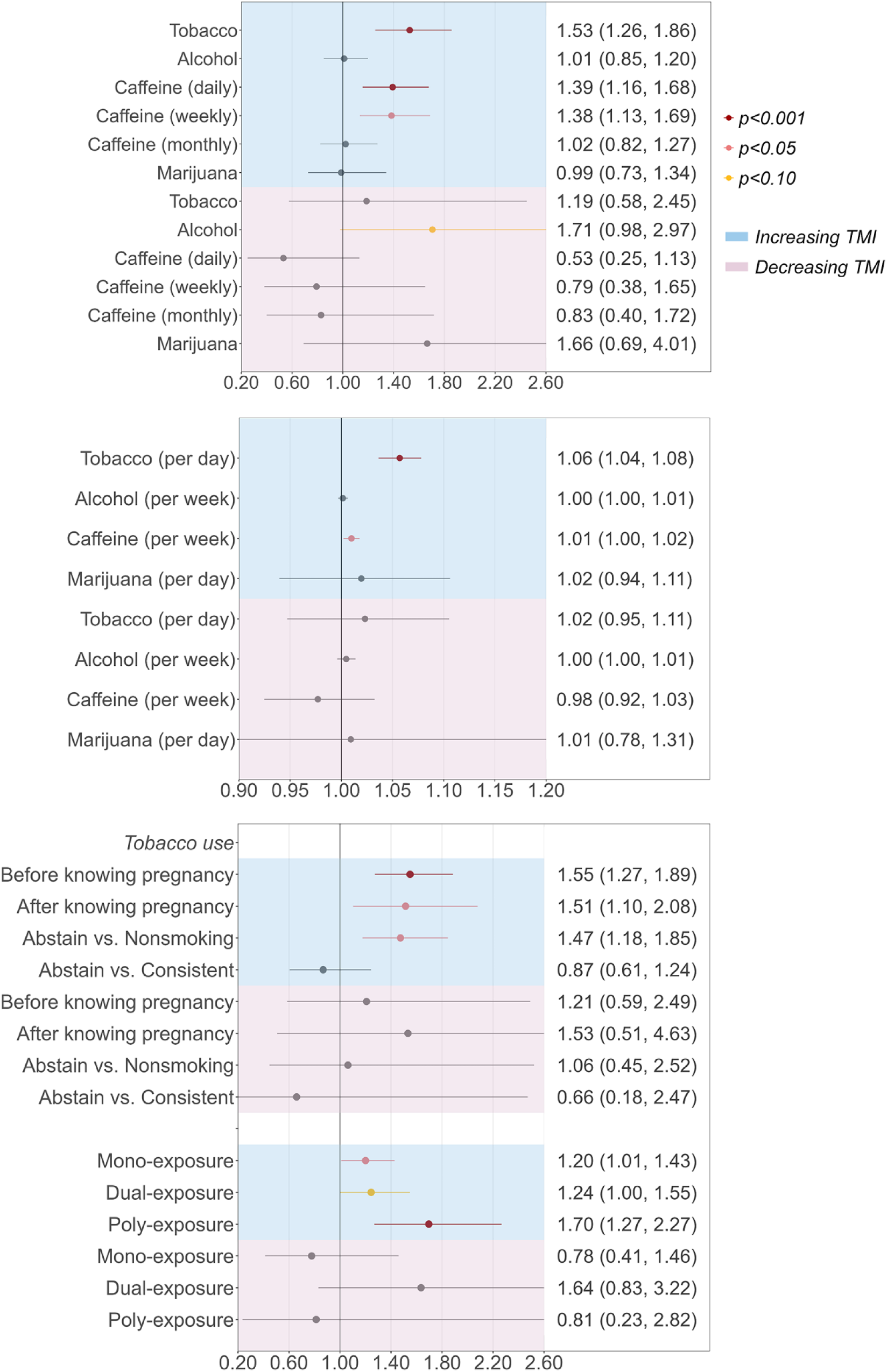
Odds ratios for prenatal substance exposure predicting TMI trajectories (reference group: Stable TMI)

## Discussion

To our knowledge, this is the first longitudinal study to explore the impact of prenatal substance exposure (PSE) on adiposity development in early adolescence, using tri-ponderal mass index (TMI) as an indicator. Latent growth mixture modeling (LGMM) identified three distinct TMI trajectories over four years: *Stable TMI*, *Increasing TMI*, and *Decreasing TMI*, highlighting subpopulations with divergent patterns of adiposity change. After controlling for various confounding factors, we found a significant effect of prenatal exposure to certain substances, particularly tobacco and caffeine, on the likelihood of children exhibiting *Increasing TMI*. These findings underscore the link between maternal substance use during pregnancy and the long-term risk of obesity in children.

The distinct TMI trajectories observed during the follow-up period highlight the variability in adiposity development among children predominantly aged 9 to 14, reflecting their transition from pre-adolescence to early adolescence. Most participants (86.6%) exhibited a *Stable TMI* trajectory, indicating constantly low and stable adiposity during this period, while 12.5% showed a consistent increase over four years. The *Decreasing TMI* trajectory (0.9%) displayed a declining pattern of TMI, yielding less stable estimates due to the small sample size. Importantly, children with *Increasing TMI* had a higher baseline TMI, suggesting that many with overweight/obesity may have begun developing these issues before adolescence. Previous research has often examined adiposity development trajectories using group-based trajectory models (e.g., LCGM), which primarily focus on differential levels between groups rather than individual variations (slope) within each group [21,22]. However, expecting more heterogeneity in TMI levels and slopes over time, we employed LGMM, with a superior model fit supporting this hypothesis.

Prenatal tobacco exposure significantly increased the likelihood of following the *Increasing TMI* trajectory rather than the *Stable TMI* trajectory, aligning with prior research linking prenatal tobacco exposure to childhood overweight [30,31]. Our study extends previous findings on the association between early tobacco exposure and later obesity by demonstrating that the effects of prenatal tobacco exposure persist into adolescence, with higher amounts of daily smoking indicating an increased risk of TMI growth during early adolescence. Several mechanisms, although not fully elucidated, have been proposed to explain this relationship, including oxidative stress, changes in placental function, alterations in central nervous system regulation of energy balance, increased sensitivity to obesogenic diets, and reduced physical activity [9,32].

Tobacco exposure both before and after awareness of pregnancy played a role in predicting an increase in TMI, underscoring the importance of early gestational exposure. Notably, mothers who quit smoking after becoming aware of their pregnancy did not show significant improvement in their children’s adiposity outcomes, compared to those who continued smoking. This suggests that tobacco exposure early in pregnancy may have lasting effects on adolescents’ adiposity development. Most previous research on smoking cessation has focused on infants or preschool children. For instance, mothers who stop smoking by the third month of pregnancy tended to have infants with similar birth weights to those of nonsmoking mothers [33]. A recent meta-analysis indicated that although early cessation during pregnancy may be beneficial compared to continued smoking, there is still a significantly higher risk of overweight and obesity in children of mothers who quit smoking during pregnancy compared to those of nonsmoking mothers, which partially supports our findings [34]. Regardless, evidence regarding adolescents remains largely scarce. Our findings suggest that the adverse effect can occur even before the early stage of pregnancy, highlighting the importance of preconception health education and early pregnancy interventions to reduce tobacco use.

The significant effect of prenatal caffeine exposure on adolescent adiposity development is noteworthy. While prior studies have linked maternal caffeine intake during pregnancy to offspring’s risk of obesity, little is known about the long-term influences [14,35]. Supporting the growing body of evidence, both daily and weekly prenatal caffeine exposure were associated with increased adiposity gain patterns in early adolescence, with a modest dose-response effect. Prenatal caffeine exposure may influence brain structure, reward sensitivity, food intake, and cognitive processing, potentially increasing the risk of obesity later in life [36,37]. Animal studies also suggest that prenatal caffeine ingestion may alter glucose metabolism and neuroendocrine function [38,39]. Our findings indicate that even moderate levels of caffeine intake during pregnancy (e.g., more than once a week but less than daily) may have lasting effects on children’s fat development, underscoring the need for more rigorous guidelines on caffeine consumption during pregnancy.

Additionally, unlike tobacco and caffeine, prenatal alcohol exposure was associated with lower TMI at a single time point, and a strong trend linking it to *Decreasing TMI* trajectory persisted even after controlling for confounders. This may, to some extent, support the notion that early alcohol exposure has a growth-restricting effect [40,41]. However, existing evidence on their relationship is inconsistent [10–12]. The discrepancies may be attributed to variations in adiposity metrics, study designs, and various confounding factors [42]. Regarding marijuana exposure, despite being less common in *Stable TMI* group, it did not significantly predict TMI changes after controlling for confounders. Although some animal studies suggest a role for the cannabinoid system in metabolic programming [43], evidence in humans remains limited and mixed, likely due to complex biological mechanisms, sex differences, and co-use of substances like tobacco [15,44].

Although not all prenatal substance exposures significantly predisposed children to an increase in TMI during early adolescence, the dose-response effect of exposure level is particularly noteworthy. Children exposed to more than two substances prenatally were found to have substantially higher odds of following an increasing adiposity trajectory. In real-world scenarios, prenatal exposure to multiple substances is common [18,45]. However, the cumulative and potential synergistic effects of multiple substance exposures on adiposity development remain largely unexplored. The result highlights the importance of considering multiple exposures in both research and clinical contexts. The limited understanding would benefit from future studies aiming to explore the underlying mechanisms of these cumulative effects.

### Implications

The current results suggest that even socially acceptable substances can have lasting effects on childhood obesity risk. The findings have important practical implications for public health and clinical practice. They reinforce the significance of preconception and prenatal health, particularly regarding substance use. Public health campaigns may need to broaden their focus beyond alcohol and illicit drugs to include tobacco and caffeine intake before and during pregnancy. Additionally, the persistence of the effect into adolescence highlights the need for early-life interventions targeted at children with known PSE, with the goal of preventing or mitigating adverse adiposity outcomes. Furthermore, addressing polysubstance exposure is particularly critical, as the cumulative effects pose a greater risk.

### Strengths and limitations

Our study has several strengths, notably the use of a large, diverse, national dataset from the ABCD Study. The sizable *Increasing TMI* group provides robust estimates for our primary focus on obesity-related outcomes. Moreover, the five-time-point modeling enables tracking adiposity development over time. Additional strengths include using TMI as a more precise and comparable metric for childhood adiposity than traditional BMI, and a careful and relatively comprehensive adjustment for confounding factors.

Some limitations of this study should be acknowledged. First, retrospective reports of PSE may introduce recall bias. Second, the small size of the *Decreasing TMI* group risks overfitting by capturing random noise, leading to unstable estimates and interpretation challenges; nevertheless, based on visual observation of individual trajectories within this group, the model appeared to be less affected by noise and captured this trend. In fact, it is reasonable that significant weight loss during adolescence is uncommon in generally healthy populations. Third, although a wide range of confounding factors were considered, there were no data on other factors that may affect offspring growth, such as maternal obesity, maternal diet, paternal substance use, and postpartum behaviors. Fourth, findings regarding the effect of prenatal caffeine exposure were limited by the absence of information on its timing. Future research would benefit from more detailed data on the dose and timing of caffeine exposure. Fifth, while TMI is a reliable indicator of body fat in adolescents, additional measures of body composition, like bioelectrical impedance analysis or dual-energy X-ray absorptiometry, would provide a more comprehensive assessment of adiposity. Finally, while this study followed adolescents over four years, extended follow-up research is clearly needed to understand the persistence of these trajectories into late adolescence and adulthood, as well as their association with long-term health outcomes.

## Conclusions

This study provides compelling evidence for the long-term role of prenatal substance exposure, particularly tobacco and caffeine, in predisposing children to increasing adiposity over four years from pre-adolescence through early adolescence. The findings provide insights into the limited understanding of determinants of body fat development in adolescence. The present study re-emphasizes the significance of early life factors for later growth and highlight the importance of a substance-free pregnancy, suggesting that interventions aimed at reducing adolescent obesity should begin as early as the prenatal period. Future research is warranted to elucidate the mechanisms underlying these associations, such as neurocognitive functions, and to develop effective interventions that mitigate the long-term impacts of prenatal substance exposure on offspring health.

## Data availability

Data are available through the National Data Archives: https://nda.nih.gov/abcd.

## Acknowledgements

Data used in the preparation of this article were obtained from the Adolescent Brain Cognitive Development (ABCD) Study (https://abcdstudy.org/), held in the NIMH Data Archive (NDA). The ABCD Study is supported by the National Institutes of Health and National Institute on Drug Abuse and additional federal partners under award numbers U01DA041022, U01DA041025, U01DA041028, U01DA041048, U01DA041089, U01DA041093, U01DA041106, U01DA041117, U01DA041120, U01DA041134, U01DA041148, U01DA041156, U01DA041174, U24DA041123, U24DA041147, U01DA050987, U01DA050988, U01DA050989, U01DA050988, U01DA051018, U01DA051037, U01DA051038, and U01DA051039. A full list of supporters is available at https://abcdstudy.org/about/federal-partners/. A listing of participating sites and a complete listing of the study investigators can be found at https://abcdstudy.org/principal-investigators/. The ABCD Study consortium investigators designed and implemented the study and/or provided data but did not necessarily participate in analysis or writing of this report. This manuscript reflects the views of the authors and may not reflect the opinions or views of the NIH or other ABCD Study consortium investigators. The ABCD Study data repository grows and changes over time. The ABCD Study data used in this report came from https://doi.org/10.15154/z563-zd24. DOIs can be found at 10.15154/jkj4-cd64.

The authors would like to sincerely thank all participants in the ABCD Study.

## Author contributions

RL and JJT were involved in conceiving and designing the study. JJT ensured access to the data. RL conducted the data analyses. IMW, IS, and AJ contributed to the interpretation of the data. JJT and IMW provided funding support for this work. RL wrote the first draft of the manuscript. All authors reviewed and approved the manuscript.

## References

1. The GBD 2015 Obesity Collaborators. Health Effects of Overweight and Obesity in 195 Countries over 25 Years. New England Journal of Medicine. 2017;377:13–27.

2. Nishtar S, Gluckman P, Armstrong T. Ending childhood obesity: a time for action. The Lancet. 2016;387:825–7.

3. Carpiniello B, Pinna F, Pillai G, Nonnoi V, Pisano E, Corrias S, et al. Obesity and psychopathology. A study of psychiatric comorbidity among patients attending a specialist obesity unit. Epidemiol Psichiatr Soc. 2009;18:119–27.

4. Llewellyn A, Simmonds M, Owen CG, Woolacott N. Childhood obesity as a predictor of morbidity in adulthood: a systematic review and meta-analysis. Obesity Reviews. 2016;17:56–67.

5. Simmonds M, Llewellyn A, Owen CG, Woolacott N. Predicting adult obesity from childhood obesity: a systematic review and meta-analysis. Obesity Reviews. 2016;17:95–107.

6. Patra J, Bakker R, Irving H, Jaddoe VWV, Malini S, Rehm J. Dose-response relationship between alcohol consumption before and during pregnancy and the risks of low birthweight, preterm birth and small for gestational age (SGA)-a systematic review and meta-analyses. BJOG. 2011;118:1411–21.

7. Behnke M, Smith VC, Committee on Substance Abuse, Committee on Fetus and Newborn. Prenatal substance abuse: short- and long-term effects on the exposed fetus. Pediatrics. 2013;131:e1009–1024.

8. Barker DJP. The origins of the developmental origins theory. J Intern Med. 2007;261:412–7.

9. Rogers JM. Smoking and pregnancy: Epigenetics and developmental origins of the metabolic syndrome. Birth Defects Res. 2019;111:1259–69.

10. Amos-Kroohs RM, Fink BA, Smith CJ, Chin L, Calcar SCV, Wozniak JR, et al. Abnormal Eating Behaviors Are Common in Children with Fetal Alcohol Spectrum Disorder. The Journal of Pediatrics. 2016;169:194–200.e1.

11. Hayes N, Reid N, Akison LK, Moritz KM. The effect of heavy prenatal alcohol exposure on adolescent body mass index and waist-to-height ratio at 12–13 years. Int J Obes. 2021;45:2118–25.

12. Carter RC, Jacobson JL, Molteno CD, Jiang H, Meintjes EM, Jacobson SW, et al. Effects of heavy prenatal alcohol exposure and iron deficiency anemia on child growth and body composition through age 9 years. Alcohol Clin Exp Res. 2012;36:1973–82.

13. Gleason JL, Sundaram R, Mitro SD, Hinkle SN, Gilman SE, Zhang C, et al. Association of Maternal Caffeine Consumption During Pregnancy With Child Growth. JAMA Netw Open. 2022;5:e2239609.

14. Li D-K, Ferber JR, Odouli R. Maternal caffeine intake during pregnancy and risk of obesity in offspring: a prospective cohort study. Int J Obes. 2015;39:658–64.

15. Moore BF. Prenatal Exposure to Cannabis: Effects on Childhood Obesity and Cardiometabolic Health. Curr Obes Rep. 2024;13:154–66.

16. Cnattingius S. The epidemiology of smoking during pregnancy: Smoking prevalence, maternal characteristics, and pregnancy outcomes. Nicotine & Tobacco Research. 2004;6:S125–40.

17. Wu T, Liao Z, Wang J, Liu M. The Accumulative Effect of Multiple Postnatal Risk Factors with the Risk of Being Overweight/Obese in Late Childhood. Nutrients. 2024;16:1536.

18. Forray A, Foster D. Substance use in the perinatal period. Curr Psychiatry Rep. 2015;17:91.

19. Peterson CM, Su H, Thomas DM, Heo M, Golnabi AH, Pietrobelli A, et al. Tri-Ponderal Mass Index vs Body Mass Index in Estimating Body Fat During Adolescence. JAMA Pediatr. 2017;171:629–36.

20. Sun J, Yang R, Zhao M, Bovet P, Xi B. Tri-Ponderal Mass Index as a Screening Tool for Identifying Body Fat and Cardiovascular Risk Factors in Children and Adolescents: A Systematic Review. Front Endocrinol. 2021;12:694681.

21. Chen Y, Dangardt F, Gelander L, Friberg P. Childhood BMI trajectories predict cardiometabolic risk and perceived stress at age 13 years: the STARS cohort. Obesity (Silver Spring). 2024;32:583–92.

22. Wang Y, Li W, Chen S, Zhang J, Liu X, Jiang J, et al. PM2.5 constituents associated with childhood obesity and larger BMI growth trajectory: A 14-year longitudinal study. Environ Int. 2024;183:108417.

23. Garavan H, Bartsch H, Conway K, Decastro A, Goldstein RZ, Heeringa S, et al. Recruiting the ABCD sample: Design considerations and procedures. Developmental Cognitive Neuroscience. 2018;32:16–22.

24. von Elm E, Altman DG, Egger M, Pocock SJ, Gøtzsche PC, Vandenbroucke JP, et al. The Strengthening the Reporting of Observational Studies in Epidemiology (STROBE) Statement: Guidelines for Reporting Observational Studies. Ann Intern Med. 2007;147:573–7.

25. Auchter AM, Hernandez Mejia M, Heyser CJ, Shilling PD, Jernigan TL, Brown SA, et al. A description of the ABCD organizational structure and communication framework. Developmental Cognitive Neuroscience. 2018;32:8–15.

26. Barch DM, Albaugh MD, Avenevoli S, Chang L, Clark DB, Glantz MD, et al. Demographic, physical and mental health assessments in the adolescent brain and cognitive development study: Rationale and description. Developmental Cognitive Neuroscience. 2018;32:55–66.

27. Berlin KS, Parra GR, Williams NA. An Introduction to Latent Variable Mixture Modeling (Part 2): Longitudinal Latent Class Growth Analysis and Growth Mixture Models. Journal of Pediatric Psychology. 2014;39:188–203.

28. Jung T, Wickrama K a. S. An Introduction to Latent Class Growth Analysis and Growth Mixture Modeling. Social and Personality Psychology Compass. 2008;2:302–17.

29. Muthen LK, Muthen B. Mplus Version 8 User’s Guide. Muthen & Muthen; 2017.

30. Oken E, Levitan E, Gillman M. Maternal smoking during pregnancy and child overweight. Int J Obes (Lond). 2008;32:201–10.

31. Rayfield S, Plugge E. Systematic review and meta-analysis of the association between maternal smoking in pregnancy and childhood overweight and obesity. J Epidemiol Community Health. 2017;71:162–73.

32. Behl M, Rao D, Aagaard K, Davidson TL, Levin ED, Slotkin TA, et al. Evaluation of the association between maternal smoking, childhood obesity, and metabolic disorders: a national toxicology program workshop review. Environ Health Perspect. 2013;121:170–80.

33. Yan J, Groothuis PA. Timing of Prenatal Smoking Cessation or Reduction and Infant Birth Weight: Evidence from the United Kingdom Millennium Cohort Study. Matern Child Health J. 2015;19:447–58.

34. Perkins J, Re T, Ong S, Niu Z, Wen X. Meta-Analysis on Associations of Timing of Maternal Smoking Cessation Before and During Pregnancy With Childhood Overweight and Obesity. Nicotine Tob Res. 2022;25:605–15.

35. Papadopoulou E, Botton J, Brantsæter A-L, Haugen M, Alexander J, Meltzer HM, et al. Maternal caffeine intake during pregnancy and childhood growth and overweight: results from a large Norwegian prospective observational cohort study. BMJ Open. 2018;8:e018895.

36. Zhang R, Manza P, Volkow ND. Prenatal caffeine exposure: association with neurodevelopmental outcomes in 9- to 11-year-old children. Journal of Child Psychology and Psychiatry. 2022;63:563–78.

37. Agarwal K, Manza P, Tejeda HA, Courville AB, Volkow ND, Joseph PV. Prenatal Caffeine Exposure Is Linked to Elevated Sugar Intake and BMI, Altered Reward Sensitivity, and Aberrant Insular Thickness in Adolescents: An ABCD Investigation. Nutrients. 2022;14:4643.

38. Liu Y, Xu D, Feng J, Kou H, Liang G, Yu H, et al. Fetal rat metabonome alteration by prenatal caffeine ingestion probably due to the increased circulatory glucocorticoid level and altered peripheral glucose and lipid metabolic pathways. Toxicology and Applied Pharmacology. 2012;262:205–16.

39. Xu D, Wu Y, Liu F, Liu YS, Shen L, Lei YY, et al. A hypothalamic–pituitary–adrenal axis-associated neuroendocrine metabolic programmed alteration in offspring rats of IUGR induced by prenatal caffeine ingestion. Toxicology and Applied Pharmacology. 2012;264:395–403.

40. Carter RC, Jacobson JL, Sokol RJ, Avison MJ, Jacobson SW. Fetal alcohol-related growth restriction from birth through young adulthood and moderating effects of maternal prepregnancy weight. Alcoholism, clinical and experimental research. 2012;37:452.

41. Edwards AC, Jacobson SW, Senekal M, Dodge NC, Molteno CD, Meintjes EM, et al. Fetal Alcohol-Related Postnatal Growth Restriction Is Independent of Infant Feeding Practices and Postnatal Alcohol Exposure in a Prospective South African Birth Cohort. Nutrients. 2023;15:2018.

42. Mattson SN, Bernes GA, Doyle LR. Fetal Alcohol Spectrum Disorders: A review of the neurobehavioral deficits associated with prenatal alcohol exposure. Alcohol Clin Exp Res. 2019;43:1046–62.

43. Patrizia C, Viviana T, Maura P, Luigia T, Vincenzo C. Developmental exposure to cannabinoids causes subtle and enduring neurofunctional alterations. International review of neurobiology. 2009;85:117–33.

44. Kong KL, Lee J, Shisler S, Thanos PK, Huestis MA, Hawk L, et al. Prenatal tobacco and cannabis co-exposure and offspring obesity development from birth to mid-childhood. Pediatric Obesity. 2023;18:e13010.

45. Tran EL, England LJ, Park Y, Denny CH, Kim SY. Systematic Review: Polysubstance Prevalence Estimates Reported during Pregnancy, US, 2009–2020. Maternal and child health journal. 2023;27:426.

