## Supplementary for "Effects of Prenatal Substance Exposure on Longitudinal Tri-Ponderal Mass Index Trajectories from Pre- to Early Adolescence in the ABCD study"

Ru Li

November 18, 2024

### 1 Sample exclusion criteria

To ensure correct model estimates, participants with measurement errors and outliers were excluded based on the following criteria: 1) children with obviously impossible measurement values (e.g., height = 6.5 inches) at any time point; 2) children who were taller at an earlier time point than at subsequent time points by more than an inch; 3) children with extremely low or high sex-specific TMI values. In this study, an extremely low TMI value was defined as greater than 1.5× interquartile range (IQR) below the first quartile. Given the positive skewness and our interest in overweight, an extremely high outlier was defined as greater than 3× IQR above the third quartile. Additionally, children with diagnosed eating disorders (previously or currently), those on medication affecting food intake (e.g., antidepressants and antipsychotics), or with mothers who reported using substances including cocaine, heroin, and oxycontin during pregnancy or lactation were excluded. Only children whose biological mothers completed the prenatal exposure questionnaire were included. Furthermore, children with mislabeled sex, incorrect sex-specific puberty, and missing data on demographics and substance use during breastfeeding were excluded, leading to a final sample of 7 881 participants across the five waves.

### 2 Background information

Past or current eating disorders at Y2 and Y4 were assessed by the Kiddie Schedule for Affective Disorders and Schizophrenia (KSADS), a semi-structured interview that includes DSM-5-based diagnostic modules for broad psychiatric disorders. Information on parent-reported medications that might affect food intake was collected at each time point.

Data on lactational exposure to tobacco, alcohol, caffeine, and marijuana were collected through a breastfeeding questionnaire and treated as confounding factors in the analyses when applicable. Information on childhood substance use during follow-ups was collected through the Substance Use Interview and coded into two categories: non-use and use (since the last meeting or in the past six months).

Parent-reported demographic information about the children, including age, sex at birth, and race divided into five categories (White, Black, Asian, Hispanic, Other), was collected at Y0. The sex-specific Pubertal Development Scale was used to estimate pubertal Tanner stage at each time point, with the five stages categorized as pre-, early-, mid-, late-, and post-puberty [1]. Puberty scores were averaged from the stages reported by both children and parents at all time points. Basic birth information, including the child’s birth weight, maternal age at childbirth, and maternal education (categorized into 29 levels as a continuous variable) as an indicator of socioeconomic status, were also treated as confounders.

### 3 Modeling procedures

The modeling was conducted using Mplus 8.3 software [2], primarily following established instructions for performing longitudinal latent variable mixture modeling [3,4]. First, a single-class growth curve model was performed to investigate the overall development of TMI. Good model fit was indicated by values of Comparative Fit Index (CFI) and Tucker-Lewis Index (TLI) > 0.9, as well as Root Mean Square Error of Approximation (RMSEA) and Standardized Root Mean Square Residual (SRMR) < 0.08 [5]. Considering the high sensitivity of the χ² statistic to sample size and the assumption of multivariate normality, its significance should not be the primary criterion for model rejection [6]. Therefore, we did not rely on the χ² test results to assess model fit.

Second, multi-class models were conducted by incrementally increasing the number of latent groups from two to five. Latent growth mixture modeling (LGMM), with within-class variances freely estimated, was conducted to explore the heterogeneity in TMI changes and to identify subpopulations with distinct TMI growth patterns. In addition, a latent class growth model (LCGM), which assumes no within-class variances, was used as a potential alternative model. For the modeling, we employed robust maximum likelihood estimation (*ANALYSIS: ESTIMATOR = MLR*) for mixture model analysis (*TYPE = MIXTURE*) with 500 random starts (*STARTS = 500 20*) and 20 initial iterations (*STITERATIONS = 20*) to ensure global optimization and enhance the stability of class-specific parameter estimation.

The optimal model was evaluated based on the following indices: higher Log-likelihood (LL) values, lower Akaike Information Criterion (AIC) and Bayesian Information Criterion (BIC) values indicating better model fit; Vuong-Lo-Mendell-Rubin Likelihood Ratio Test (LMR), Bootstrapping Likelihood Ratio Test (BLRT) for k versus k-1 groups, where p-values below 0.05 suggest that k groups have significant better fit than k-1 groups [7]. Moreover, entropy values and Average Latent Class Probabilities (ALCP) greater than 0.80 indicate good classification accuracy. We also considered the Most Likely Latent Class Membership (MLLC) and aimed to avoid small sample size within a group (e.g., MLLC < 1%).

Finally, parameter estimates from the two seed values corresponding to the best log-likelihood values were then examined, with replicated estimates ensuring no issues related to non-convergence or local maxima. To determine the reliability and robustness of the modeling, a sensitivity analysis was performed.

### 4 Attrition analysis for missing data mechanism

In the modeling process, missing data on TMI were addressed using full information maximum likelihood (FIML). FIML preferably assumes that missing data are either missing at random (MAR) or completely at random (MCAR). Hence, to justify the reliability of using FIML, the missing data mechanism was analyzed.

Logistic regression analyses were conducted with the missingness of TMI at each time point as the outcome variable and sample characteristics as predictors. The missingness of TMI at Y0 was minimal and not associated with any variables. The relationships between sample characteristics and TMI missingness varied across time points, with each variable exhibiting inconsistent positive, negative, or non-significant correlations, indicating a more random missing data mechanism. This variability in relationships supports the idea that missing data occurred due to random factors rather than systematic biases. Additionally, when comparing TMI differences between groups with and without missing data across five time points, only the last time point (p < .001) showed a significant difference, with the missing group having higher TMI. For the first four time points, there were no significant differences in TMI (p = .658, .696, .273, and .276 for Y0, Y1, Y2, and Y3, respectively). Overall, it is reasonable to assume that the missing data mechanism is random.

### 5 Detailed results of modeling

#### 5.1 Growth curve modeling

We conducted a linear and quadratic growth curve model. As presented in **Table S1**, quadratic modeling showed a better overall fit to the data. **Figure S1** depicts the single-trajectory model and observed individual trajectories. The overall TMI did not significantly change over time. However, there were significant variances in baseline level and changes, which suggested individual differences in the TMI development. Additionally, measurement error variance was constrained to be equal across all time points. The constrained model with equal error variances in time was also supported by a good model fit (**Table S1**), indicating stable measurement reliability over time. The model results ensured the robustness and interpretability of subsequent modeling to identify distinct growth patterns among subgroups.

**Table S1.** Fit indices of single-class growth curve modeling

|  | **CFI** | **TLI** | **RMSEA** | **SRMR** | **AIC** | **BIC** | **L (Var)** | **S (Var)** | **Q (Var)** |
| --- | --- | --- | --- | --- | --- | --- | --- | --- | --- |
| **Linear** | 0.991 | 0.991 | 0.052 | 0.032 | 39992.0 | 40061.7 | -0.043^***^ (0.447)^***^ | 0.001 (0.014)^***^ |  |
| **Quadratic** | 0.997 | 0.995 | 0.037 | 0.022 | 39847.0 | 39944.6 | -0.042^***^ (0.455)^***^ | -0.004 (0.058)^***^ | 0.001 (0.003)^***^ |
| **Constrained** | 0.987 | 0.987 | 0.063 | 0.030 | 40093.2 | 40162.9 | -0.040^***^ (0.420)^***^ | -0.006 (0.021)^***^ | 0.001 (0.002)^***^ |

Constrained: Measurement error variance equal across all time points.

CFI: Comparative Fix Index, TLI: Tucker-Lewis Index, RMSEA: Root Mean Square Error of Approximation, SRMR: Standardized Root Mean Square Residual, AIC = Akaike Information Criterion, BIC = Bayesian Information Criterion.

L (Var) = Level (variance), S (Var) = Linear slope (variance), Q (Var) = Quadratic slope (variance).

^***^ P < .001

**Figure S1.** Single-trajectory based on growth curve model

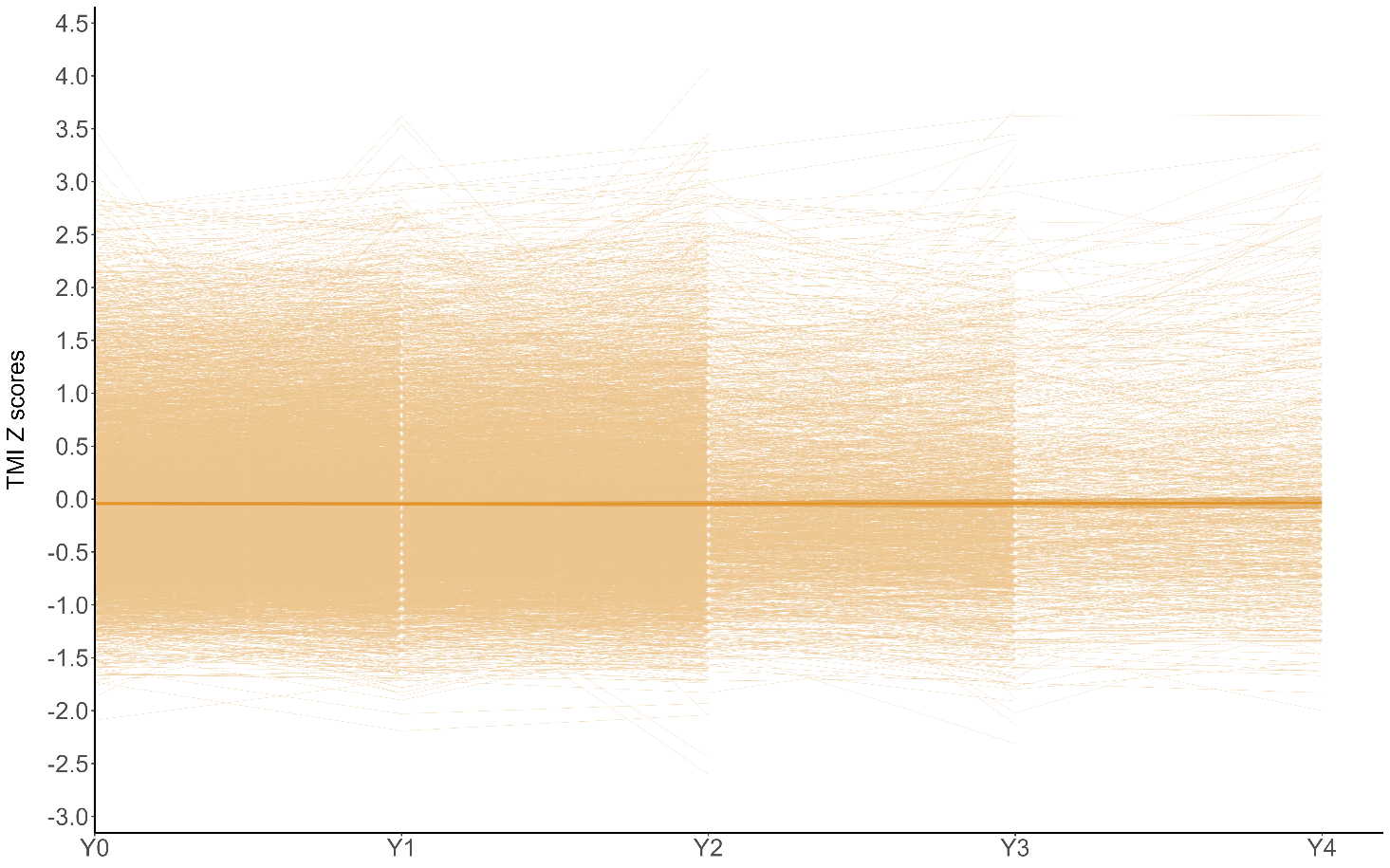

#### 5.2 Latent growth mixture modeling (LGMM)

After testing various model specifications, we selected a quadratic model with freely estimated intercepts across latent classes. However, based on the model specification information, linear and quadratic slopes cannot be freely estimated across latent classes; therefore, they were fixed to be equal, reflecting variances within individual groups while remaining invariant between groups.

An optimal model was specified, with indices for determining the number of latent classes presented in **Table S2**. The quadratic model exhibited a better model fit than the linear model. The best model could not be identified up to the 5-class solutions based on the increasing LL value, the decreasing AIC and BIC values, and entropy values greater than 0.7. However, the LMR test suggested no significant improvement when the number increased up to three classes. Moreover, compared to the 3-class model, the 4-class and 5-class models exhibited a more skewed class distribution due to the small sample sizes in more than one latent class. Moreover, the very small classes (with MLLC < 1%) pose a risk of overfitting, as they may capture random noise rather than meaningful patterns, leading to less stable parameter estimates and making it more difficult to interpret and apply practically. This was supported by the trajectory plot, which showed that these small classes were primarily mixtures of the larger classes. Accordingly, the 3-class solution was preferred for its better balance between fit and interpretability. Replicated estimates using the two seed values with the best log-likelihood values showed no issues of non-convergence or getting stuck in local maxima.

**Table S2.** Indices for model selection (latent growth mixture modeling)

|  |  | **Number of Latent Class** | | | |
| --- | --- | --- | --- | --- | --- |
|  |  | **2** | **3** | **4** | **5** |
| **Linear** |  |  |  |  |  |
| Model Fit | *LL* | -19447.8 | -18967.9 | -18374.7 | -18200.4 |
|  | *AIC* | 38923.6 | 37971.7 | 36793.4 | 36452.9 |
|  | *BIC* | 39021.2 | 38097.2 | 36946.7 | 36634.2 |
|  | *LMR (p)* | < .001 | < .001 | < .001 | .026 |
|  | *BLRT* | < .001 | < .001 | < .001 | < .001 |
| Classification  Quality | *Entropy* | 0.85 | 0.91 | 0.92 | 0.92 |
|  | *MLLC (%)* | 88.9/11.1 | 88.7/10.2/1.2 | 87.5/10.3/1.1/1.0 | 86.6/8.9/2.8/1.2/0.5 |
|  | *ALCP* | 0.97/.85 | .97/.85/.94 | .97/.86/.93/.92 | .97/.81/.80/.91/.97 |
| **Quadratic** |  |  |  |  |  |
| Model Fit | *LL* | -19176.6 | -18543.4 | -18146.8 | -17892.2 |
|  | *AIC* | 38395.3 | 37142.7 | 36363.6 | 35868.4 |
|  | *BIC* | 38541.7 | 37337.9 | 36607.7 | 36161.2 |
|  | *LMR (p)* | < .001 | < .001 | .058 | .124 |
|  | *BLRT* | < .001 | < .001 | < .001 | < .001 |
| Classification  Quality | *Entropy* | 0.99 | 0.86 | 0.87 | 0.77 |
|  | *MLLC (%)* | 99.2/0.8 | 86.6/12.5/0.9 | 85.0/12.0/2.1/0.9 | 80.3/13.5/3.2/2.1/0.9 |
|  | *ALCP* | 1.00/0.95 | .95/.85/.94 | .95/.84/.86/.95 | .89/.73/.78/.85/.93 |

LL = Log-likelihood, AIC = Akaike Information Criterion, BIC = Bayesian Information Criterion, LMR = Vuong-Lo- Mendell-Rubin Likelihood Ratio Test, BLRT = Bootstrapping Likelihood Ratio Test, MLLC = Most Likely Latent Class Membership, ALCP = Average Latent Class Probabilities.

The latent class growth modeling (LCGM) assuming within-class invariances was also considered as a potential alternative model specification. The LCGM yielded a 5-class solution as the optimal model. It showed a significantly lower log-likelihood value (-19 987.5, p < .001) and higher AIC (40 024.9) and BIC (40 199.3) compared to LGMM. This indicated that the LGMM provided a better fit for the data, making it the optimal approach for modeling the TMI trajectories in this study.

### 6 Sensitivity analysis

To assess robustness of modeling, three models with different sample sizes were compared: the specified optimal model, one model with complete data from the smallest sample size (Y3, N = 2 596), and one with less strict criteria for sample exclusion (excluded only cases with measurement errors and extreme values N = 11 124). The trajectory patterns from these models were largely consistent (**Figure S2**), supporting the reliability of FIML and the model’s robustness against biases from missing data and sample exclusions.

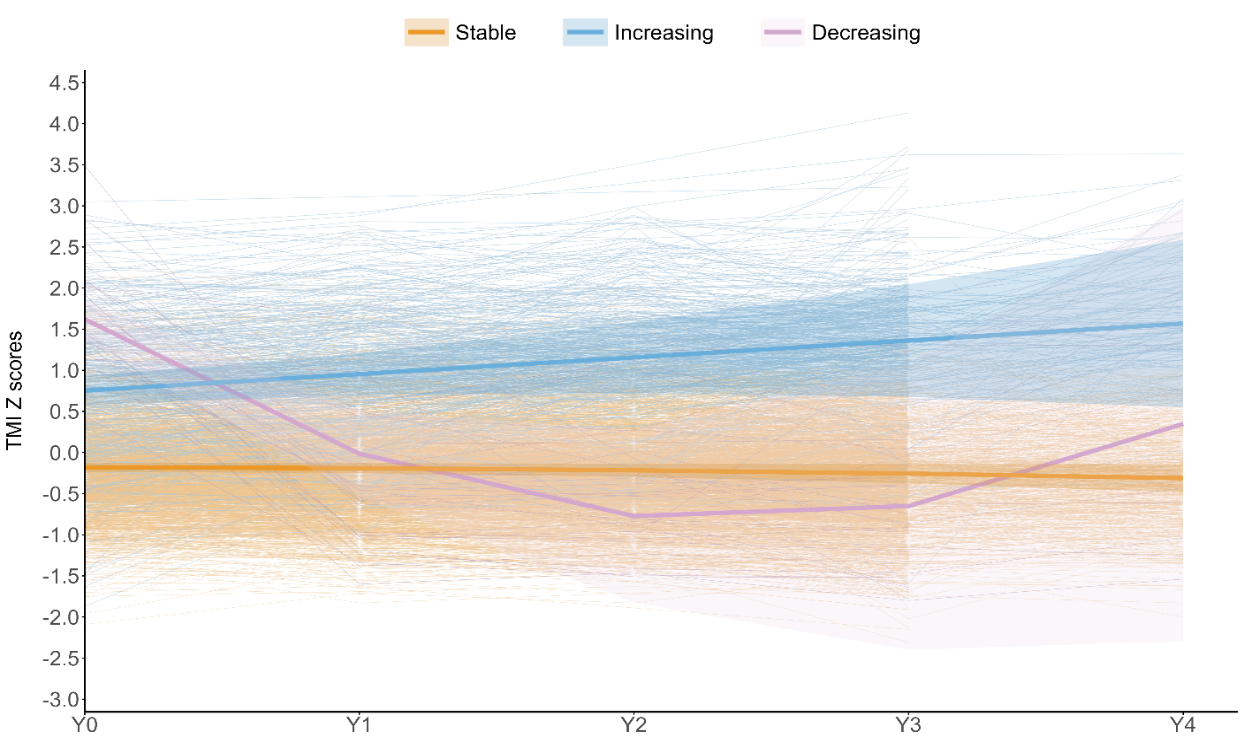

**A**

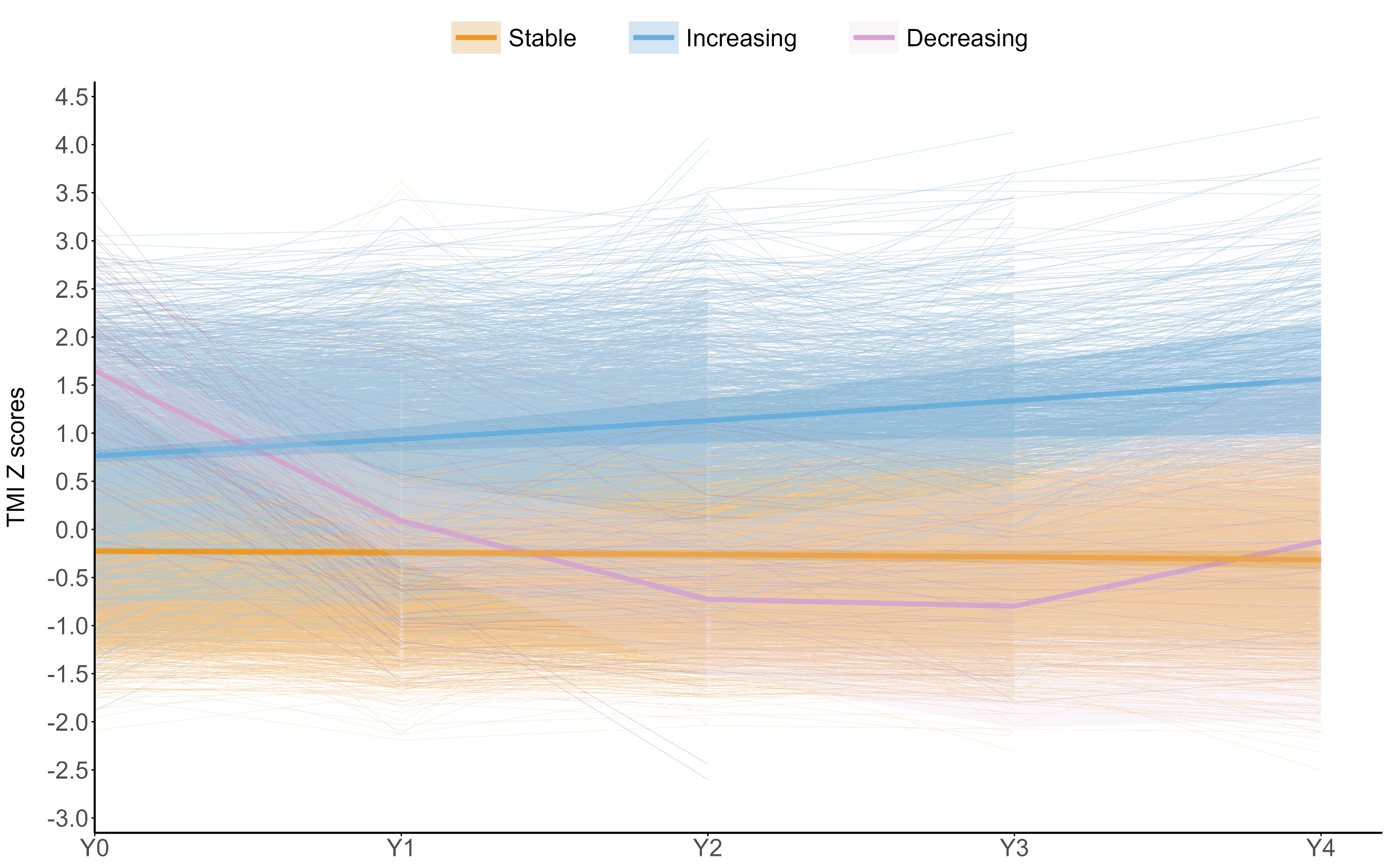

**B**

**Figure S2.** Latent growth mixture modeling with different sample sizes (A: N = 2596; B: N = 11124)

**Figure S3.** Point-biserial coefficient for correlation between prenatal substance exposure and single-time-point TMI

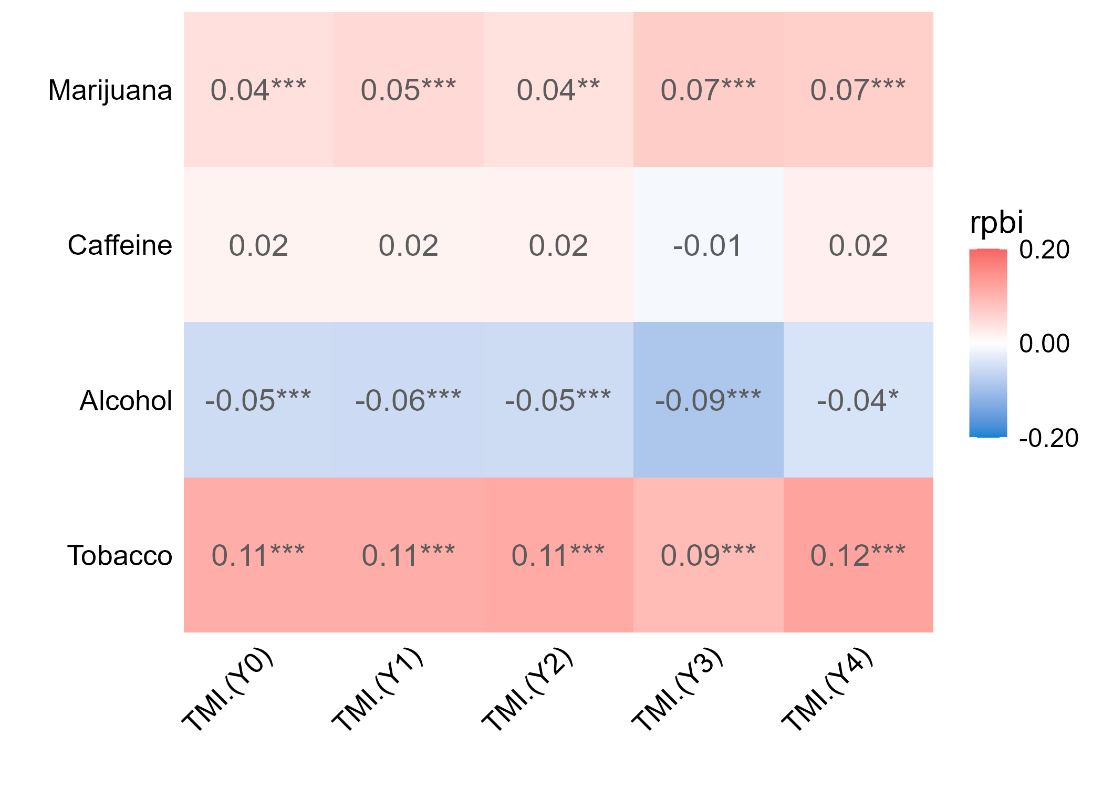

^*^*P* < 0.05, ^***^*P* < 0.01, ^***^*P* < 0.001

**References**

1. Petersen AC, Crockett L, Richards M, Boxer A. A self-report measure of pubertal status: Reliability, validity, and initial norms. J Youth Adolescence. 1988;17:117–33.

2. Muthen LK, Muthen B. Mplus Version 8 User’s Guide. Muthen & Muthen; 2017.

3. Berlin KS, Parra GR, Williams NA. An Introduction to Latent Variable Mixture Modeling (Part 2): Longitudinal Latent Class Growth Analysis and Growth Mixture Models. Journal of Pediatric Psychology. 2014;39:188–203.

4. Jung T, Wickrama K a. S. An Introduction to Latent Class Growth Analysis and Growth Mixture Modeling. Social and Personality Psychology Compass. 2008;2:302–17.

5. Ullman JB. Structural Equation Modeling: Reviewing the Basics and Moving Forward. Journal of Personality Assessment. 2006;87:35–50.

6. Schermelleh-Engel K, Moosbrugger H, Müller H. Evaluating the Fit of Structural Equation Models: Tests of Significance and Descriptive Goodness-of-Fit Measures. Methods of Psychological Research. 2003;8:23–74.

7. Ram N, Grimm KJ. Methods and Measures: Growth mixture modeling: A method for identifying differences in longitudinal change among unobserved groups. International Journal of Behavioral Development. 2009;33:565–76.
